# The impact of involving young children in improving the understanding of a new eye test

**DOI:** 10.1101/19001362

**Authors:** Therese Casanova, Carla Black, Sheima Rafiq, Jessica Hugill, Jenny C A Read, Kathleen Vancleef

**Affiliations:** NHS Business Services Authority, Stella House, Goldcrest Way, Newcastle, NE5 8NY; Pennine Care NHS Foundation Trust, Children’s Acute and Ongoing Needs Service, Callaghan House, Cross Street, OL10 2DY, Heywood, United Kingdom; Durham University, School of Education, DH1 1TA, Durham, United Kingdom; University of Oxford, Department of Experimental Psychology, Anna Watts Building, Radcliffe Observatory Quarter, Woodstock Road, University of Oxford, OX2 6GG, United Kingdom; Newcastle University, Institute of Neuroscience, Henry Wellcome Building, Framlington Place, NE2 4HH, Newcastle-upon-Tyne, United Kingdom

**Keywords:** patient and public involvement, stereotest, children, test development, PPI, med tech, stereopsis, engagement, co-production

## Abstract

**Background:** Although considered important, the involvement of young children in research is rare and its impact has never been measured. We aim to provide a good practice example of involvement of young children in the development of a tablet-based 3D eye test or stereotest, ASTEROID. We focus on improving understanding of test instructions.

**Methods:** After a pre-measure of understanding was taken, we explored issues with the test instructions in patient and public involvement (PPI) sessions. Feedback was collected via observations, rating scales and children’s comments. An interdisciplinary panel reviewed the feedback and decided on the implementation. Subsequently, a post-measure was collected. In Study 1, 650 children (2-11.8 years old) took part in the pre-measure, 111 children (1-12 years old) in the PPI sessions, and 52 children (4-6 years old) in the post-measure which also served as a pre-measure for Study 2. PPI sessions for Study 2 attracted 122 children (1-12 years old) and adults. 53 people (2-39 years old) participated in the post-measure of Study 2.

**Results:** Following feedback in Study 1, we added a frame cue and included a shuffle animation. This increased the percentage of correct practice trials from 76% to 97% (t(231)=14.29, p<.001). After adding a cardboard demo in Study 2, the percentage of correct trials remained stable but the number of additional instructions given decreased (t(103)=3.72, p<.001).

**Conclusions:** Our study demonstrates impact of involvement of very young children in research through accessible activities. It can encourage researchers to involve young children and contribute to developing guidelines.

## Introduction

The UK National Advisory Group promoting public involvement in health and social care research, INVOLVE, defines Patient and Public Involvement (PPI) as research which is …”carried out ‘with’ or ‘by’ members of the public rather than ‘to’, ‘about’ or ‘for’ them ^1^. This does not include people in their role as research subjects or participants in a study, but rather as advisers or collaborators in the research ^2^. PPI continues to be of growing importance for research across disciplines. Involvement of children in research is encouraged in the guiding principles of the United Nation’s convention on the Right of the Child. Article 12 acknowledges that children and young people have the right to be involved in decisions that affect their life, express their views and have their views listened to ^3^. Involving children in research from design to dissemination is therefore no longer a preferred approach but a requirement ^4^.

Traditionally, children’s perspectives have been filtered through interpretations of parents and carers rather than children being involved themselves with their unique insights into their own reality ^5^. Despite the change in policy and vision, examples of involvement of children in research are very limited, especially across the range of healthcare provision ^4,6,7^.

For instance, on 28^th^ April 2019, the INVOLVE evidence library ^8^ included 321 works of involvement of people in designing research in the health sector, 19 of which relate to children’s research and PPI. Of these 19, only seven describe original research that involves children, and none outline involvement of children under 6 years old. Dunn and colleagues ^4^ concluded that despite many “efforts to include children’s voices, translation into research and pedagogical practice is still evolving”.

Whilst securing involvement from adults may be more straightforward, involving children in research is challenging. There is the intrinsic issue of generational divide and power differences between researchers and children they endeavour to work with ^4^. Specifically, younger children are arguably at more risk of difficulty with understanding questions that ask for their feedback. The difficulty of choosing age-appropriate involvement methods for young children has been seen as a barrier to involvement ^4^ and good practice methodology on how PPI with children should be achieved is lacking ^2^. Some guidelines are available ^5,9^ but they only advise on running discussion groups with young people, generally above 12 years old. In sum, while toolkits and case studies from national bodies provide starting points from which research groups could *promote* PPI practice, a large evidence base containing research that implements this does not yet exist.

Furthermore, no evidence is available on the impact of PPI with young children upon research to drive the development of standards of good practice ^6^. Taken together, there is an urgent need for evidence on *how* young children can be involved in research and *what* impact can be achieved.

## Aims

The current paper describes how children were involved in improving children’s understanding of ASTEROID, a new 3D eye test, after initial prototype development. The purpose is to highlight the importance and impact of involving young children in the development of medical technology. Most crucially, this paper aims to contribute to the slowly growing pool of research that involves children and young people and provide evidence of its impact on research. Such an evidence base is the all-important next step to ensure that Public and Patient Involvement becomes a fundamental part of all kinds of research.

## General methods

### The ASTEROID study

Stereopsis, stereoscopic or simply stereo vision all refer to the perception of depth via binocular vision, or vision using both eyes. Normal stereoscopic vision is associated with correct development of visual functions and alignment of the eyes ^10–12^. Measuring stereopsis is therefore common in children with suspected amblyopia or strabismus. However, current ways of measuring stereopsis can be unengaging and under-sensitive (e.g. Huynh, Ojaimi, Robaei, Rose, & Mitchell, 2005). To address this, a new test, known as “Accurate STEReotest On a mobile Device” or ASTEROID, was developed on a glasses-free 3D tablet (see Figure 1 and Study 1 Methods for a detailed description, see also ^14^). The 3D tablet eradicates the need for children to wear 3D glasses during testing, something that has proven to be an issue with younger children with the currently available stereotests ^15^. To engage children and young people, ASTEROID took the form of an animated game. ASTEROID was validated against a gold standard stereotest and normative data were collected. Anecdotal evidence and feedback gathered during data collection highlighted some issues. Key to this is ensuring that each participant understood what “seeing the 3D image” would be like to ensure that any failures were genuinely due to problems with stereo vision rather than understanding test instructions. These initial outcomes triggered an iterative PPI approach.

**Figure 1.**
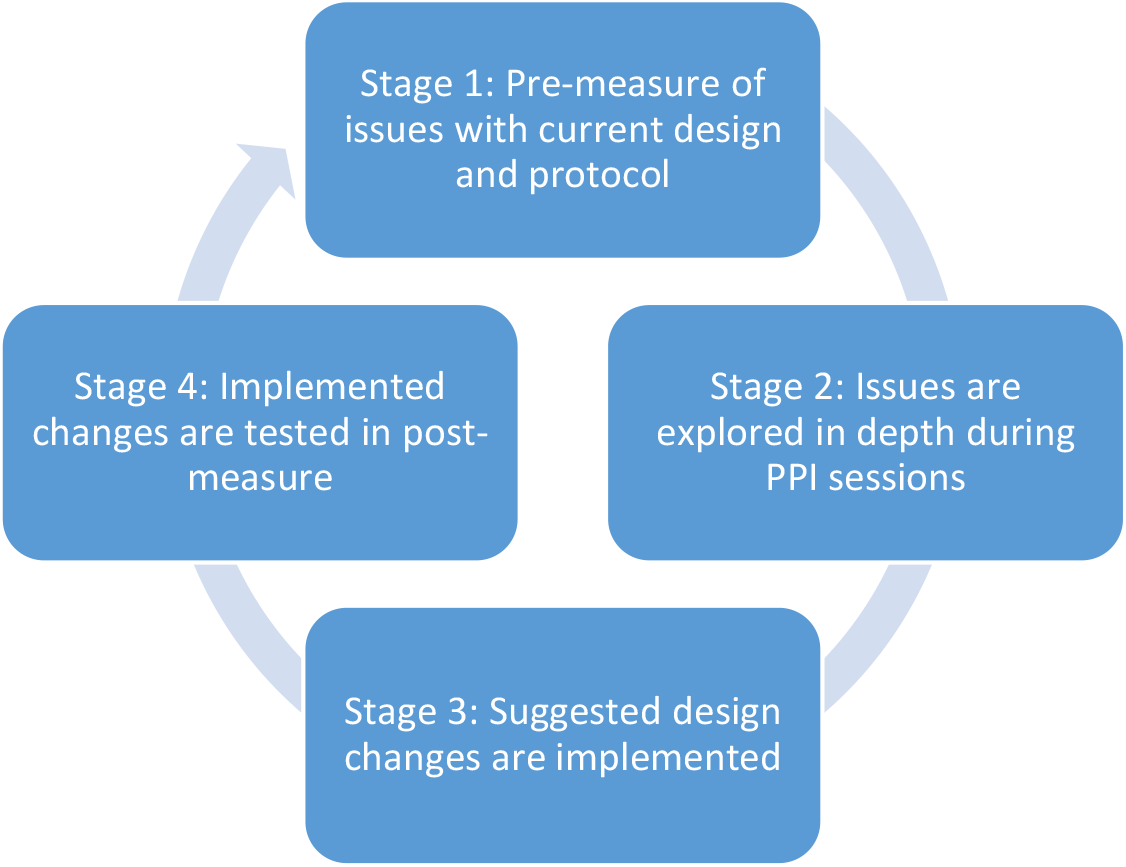
Schematic representation of how Patient and Public Involvement was embedded in the development of ASTEROID.

### Design of the studies

An iterative approach was undertaken to improve ASTEROID (see Figure 1). Each Study started with a data collection stage in which children completed ASTEROID in the context of our larger validation study and we observed an issue related to poor understanding of ASTEROID (pre-measure). In the second stage, this issue was then explored in depth during patient and public involvement (PPI) with a different group of children or adults. By collecting feedback from participants with no previous experience of ASTEROID, we were able to avoid learning effects. PPI outcome triggered changes to ASTEROID (Stage 3). Subsequently, the new version of ASTEROID was evaluated in a separate sample (post-measure, Stage 4). This enabled us to collect quantitative evidence of the impact of our PPI. Verbal feedback and observations during Stage 4 highlighted other issues and subsequently triggered another cycle. In this paper we describe two such cycles that aimed to improve understanding of ASTEROID (Study 1 and 2). Our secondary aim was to monitor how engagement levels changed following improvement of ASTEROID (Study 3).

## Study 1: Practice trials and trial-to-trial transition

### Stage 1: Pre-measure

#### Participants

650 children between 2.1 and 11.8 years old (median age = 6.2, IOR = 3.1, 318 boys and 332 girls) participated in Stage 1. They completed the ASTEROID stereotest in the context of a larger validation study with other vision tests. All parents received an information leaflet about the study and an opt-out consent form. If requested by the school or nursery an opt-in consent procedure was used. Children were always asked for oral or non-verbal assent at the time of testing. The study protocol was compliant with the Declaration of Helsinki and was approved by the Ethics Committee of the Newcastle University Faculty of Medical Sciences (approval number 01078). Data were collected in local schools and nurseries.

#### Instruments

ASTEROID (version 0.932 and 0.933) is a stereotest that runs on a 3D tablet computer with an autostereoscopic display, meaning no 3D glasses were required for use. The stereotest is embedded in an enjoyable game designed to keep children engaged and responsive. A Bayesian staircase was implemented to adjust the difficulty level of each trial, depending on the user’s ability. The test takes the form of four dynamic random-dot stereograms (Figure 2). A disparate target appears randomly in one of these stereograms. The child is verbally instructed to look at each of the four “squares” individually to see which one sticks out. The viewing distance is monitored by the front camera. To ensure that the child understands the task, ASTEROID starts with non-stereo practice trials. In these trials, a colour is added on top of the stereo cue, to make the target clearly visible. This cue is also visible even to colourblind individuals, due to the change in luminance. If the child solves the practice trials correctly, the colour cue gradually fades away over the next few trials and will eventually disappear, leaving only the stereo cue. There are a minimum of 25 stereo trials – the number of non-stereo practice trials increases with the number of mistakes made in the practice trials. Technical details of ASTEROID are described elsewhere ^14^.

**Figure 2:**
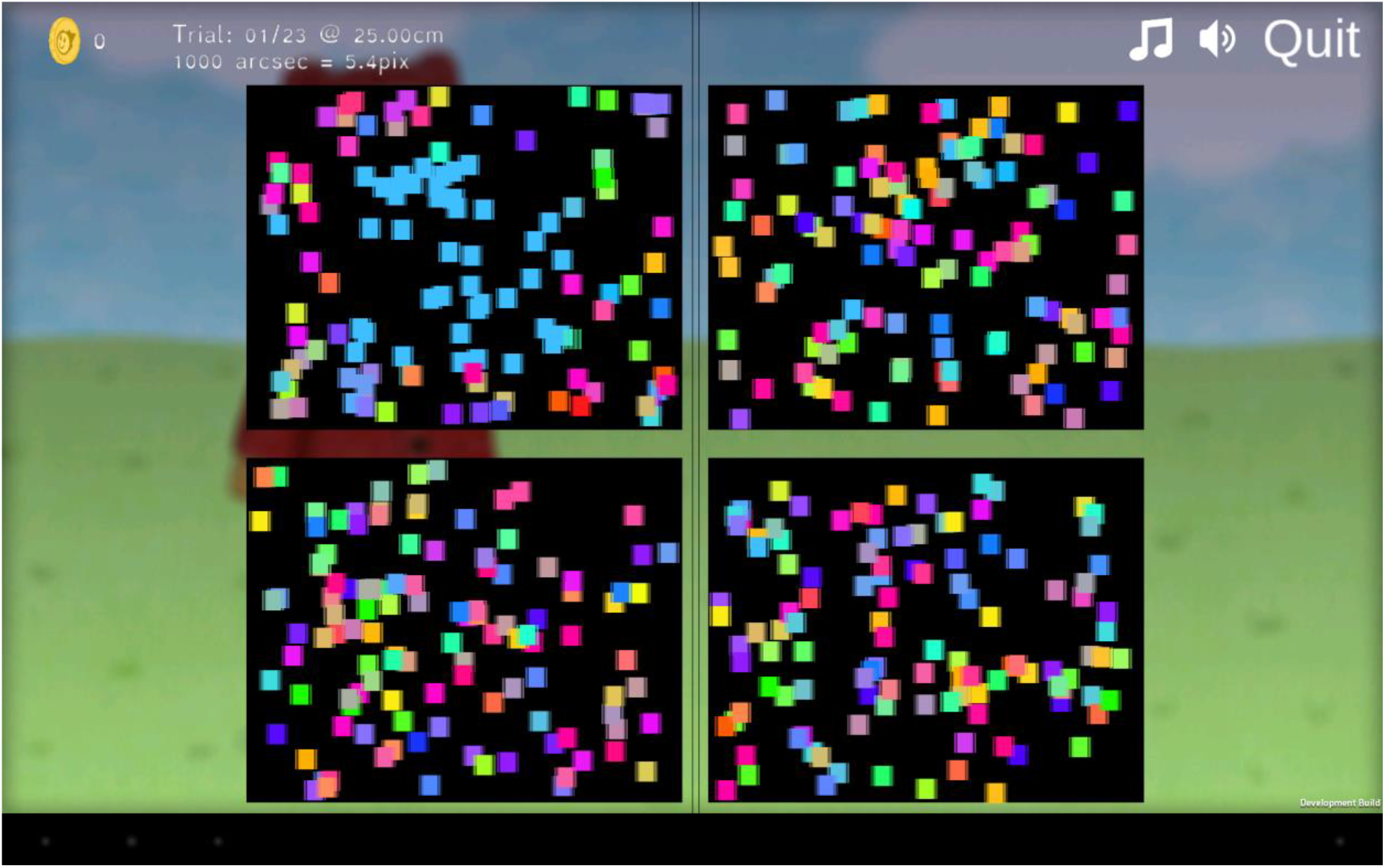
Screenshot of a practice trial of ASTEROID (version 0.933). The tests show four dynamic random-dot stereograms. One of the four stereograms has a square with a different disparity and appears to float above the display. In the practice trials this square also has different coloured dots. The colour cue is removed in the disparity-only trials.

#### Outcome measures

Our primary outcome measures were (1) the proportion of children who solved less than 80% of the practice trials, (2) the proportion of practice trials that were solved correctly by each child, and (3) the number of additional verbal instructions needed for each child. Performance in the practice trials gives us a good indication of the understanding of the task because even children with vision problems like stereoblindness or colourblindness should be able to solve the practice trials correctly. Secondary, observations of children playing ASTEROID were valuable in generating hypotheses for poor understanding of the task that were further explored in Stage 2.

#### Results

39% of the children showed a poor understanding of the ASTEROID task by solving less than 80% of the practice trials correctly. The average proportion of correct practice trials was 0.76 and on average 10 additional prompts were needed per child to explain the task. We particularly observed difficulties in the transition between the practice trials and the stereo trials.

### Stage 2: PPI sessions

#### People involved

111 children between 2 and 12 years old (median age = 4, IQR = 3, 53 girls, 55 boys, 3 gender not recorded) were involved in the PPI sessions. They attended local science and history museums in Newcastle-upon-Tyne (United Kingdom) between October 2016 and January 2017 and were invited to join a PPI session for 5-10 minutes. No ethical approval was required for PPI ^16^. Children were thanked for their feedback with a ‘Junior Scientist’ certificate and a sticker.

#### Level and nature of involvement

We organised three informal drop-in PPI sessions (Table 1). We set-up a stall with engagement activities around vision testing and neuroscience in an area of the museum with high traffic (Figure 3). The stall contained games including visual distortion goggles, bean-bags and examples of visual illusions. The competitive nature of some of the activities engaged children and their siblings, whilst certificates and other small prizes were available as a reward. Crucially, children were invited to try out ASTEROID (version 0.932 and 0.933) and comment on any aspect of the design. Opinions were gathered by research assistants: CB, TC, JH, and SR. CB and SR had experience in engaging children for vision testing in their previous roles as an optometrist and orthoptist respectively. TC has a postgraduate degree in Health Psychology and had worked with in a primary school as a Teacher Training Programme Coordinator. JH worked as a primary school teacher and completed an MSc in Psychology before joining the research team.

**Table 1.** Overview of PPI sessions for Study 1

| Session number | Date | Venue | Nature of session | Age range | N | ASTEROID version number | Individual feedback provided? | Engagement rating provided? |
| --- | --- | --- | --- | --- | --- | --- | --- | --- |
| 1 | 28/10/2016 | Discovery Museum<sup>1</sup> | Drop-in session | 3-12 | 26 | 0.932 | N | Y |
| 2 | 07/01/2017 | Great North Museum: Hancock<sup>1</sup> | Drop-in session | 2-11 | 38 | 0.933 | N | Y |
| 3 | 24/01/2017 | Centre for Life<sup>2</sup> | Drop-in session | 1-6 | 47 | 0.933 | Y | Y |

<sup>1</sup> Local museum with free entrance; <sup>2</sup> Local museum with entrance fee; N = number of people involved

**Figure 3.**
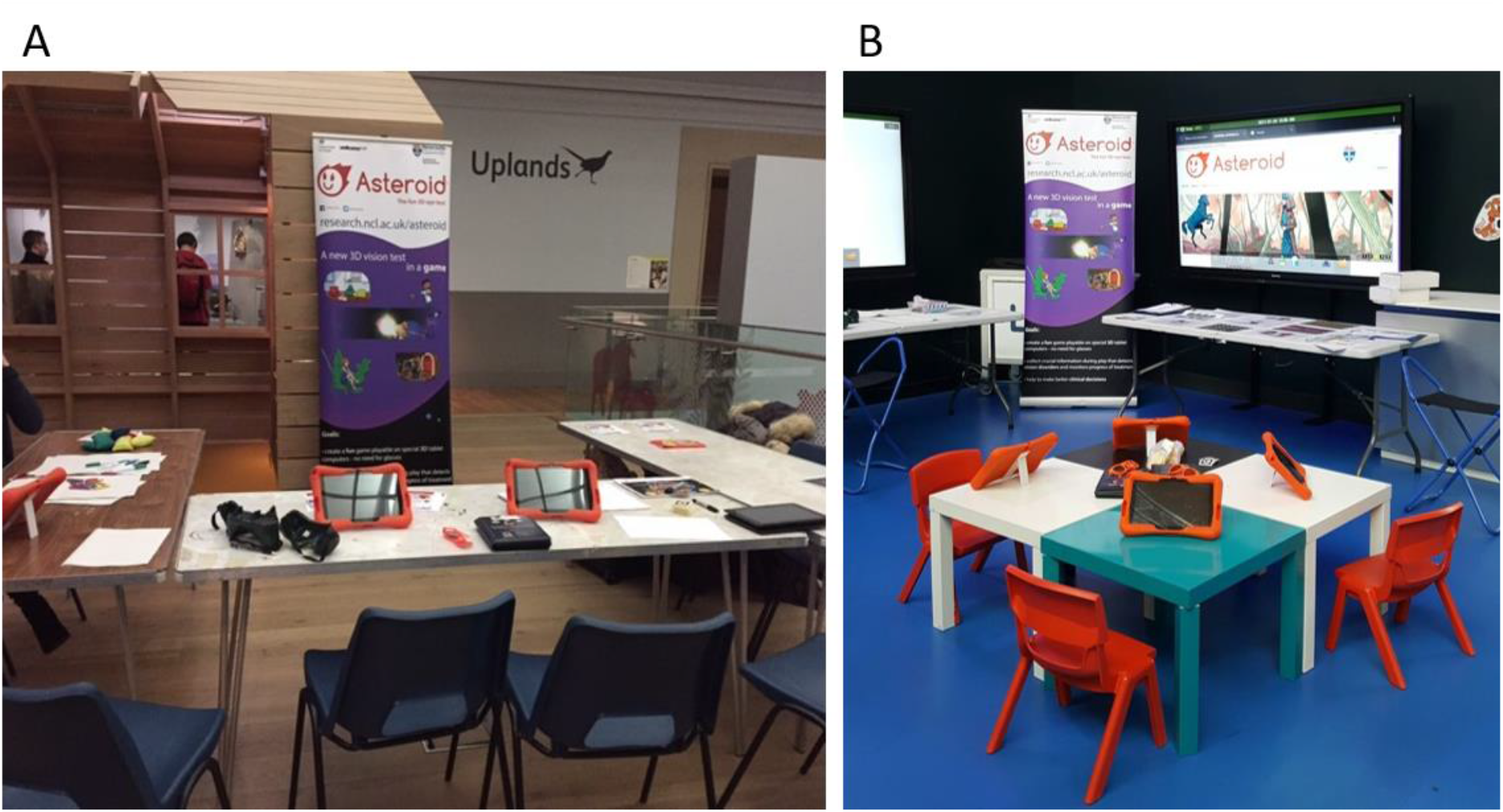
Our set-up for the drop-in PPI activities at Discovery museum (A) and Centre for Life (B)

#### Outcome measures

1. Observations of the participant’s progress (e.g. focus, hesitation, difficulties etc.) were collated on each trial.
2. Individual verbal feedback from participants: We asked questions like what they found difficult, what was unclear to them, whether they enjoyed ASTEROID, etc. People were free to give any feedback. This information was collected in Session 3 only.

#### Outcomes of PPI

Observations of some children indicated a lack of understanding of the gameplay. For example, for a 2-year old boy it was noted that he “only played a few trials and showed no understanding of what to do”. A similar observation was made for another 2-year old girl: “Happy to play but didn’t understand”, and for a 1.5 year old girl: “Didn’t understand, mum modelled a lot”. In many children we observed hesitation on how to proceed at the end of the non-stereo practice trials, indicating a gap in understanding on what to do during the non-stereo and stereo stages of the game. In addition, comments made during sessions indicated that the colour cue in combination with the colourful dots in the non-stereo trials primed children to look for a local colour change in a few dots instead of an overall depth change in the stereo trials.

Second, some children had difficulties understanding that on each trial the locations of the target was randomly determined. In subsequent trials, these children would tap all four stereograms in alternation. For instance, if they incorrectly tapped the top left stereogram in one trial, in the next trial they would not revisit this location but instead try for instance the top right location. If the target was not found there, they would move to yet another position in the third trial. This seemed to indicate that children did not understand that the placement was random, but imagined that the target location remained the same until it was found.

### Stage 3: Implementing changes

#### Methods

Feedback from Stage 2 was summarised by research assistants and discussed at a cross-disciplinary meeting attended by vision scientists, computer scientists and game developers. Once consensus was reached, the proposed changes were implemented in a new version of ASTEROID.

#### Results

To solve the first problem, we replaced the colour cue by a frame (Figure 4). The second problem was solved by adding a card shuffle animation. At the end of the trial, the stereograms would flip around, move to the centre of the screen, mix up, move out again to the four corners of the display and flip back face up (see video clip at https://youtu.be/w8q-4uejwdk). The animations mimicked playing cards being shuffled and dealt out. We believed this would explain to young children that in the next trial the target could appear in any of the four locations.

**Figure 4.**
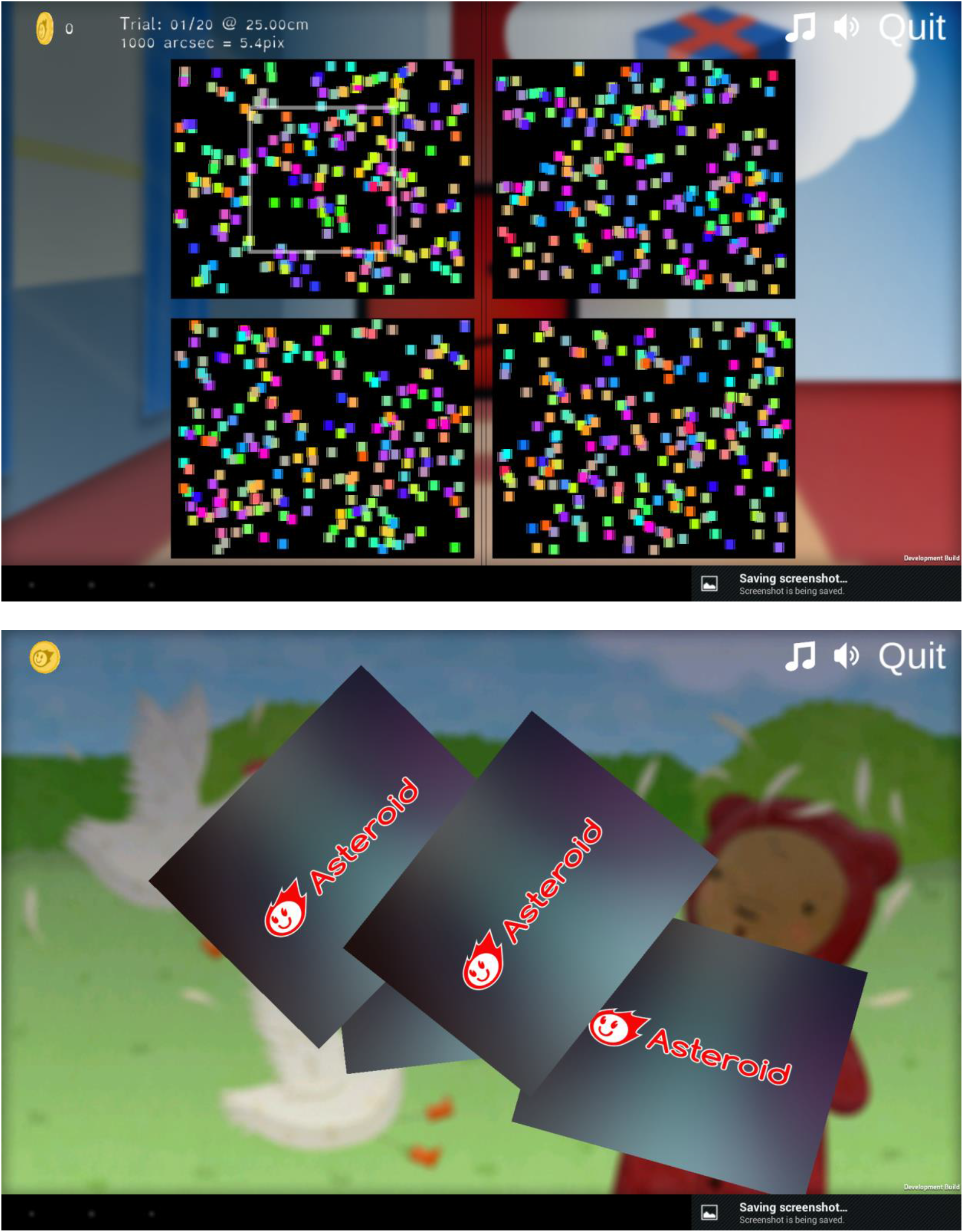
Screenshots (version 0.937) of the changes made after the first Public and Patient Involvement round: a frame cue (top) and a shuffle animation (bottom, see also video at https://youtu.be/w8q-4uejwdk).

### Stage 4: Post-measure

#### Participants

52 children between 4.5 and 6 years old (median age = 5, IQR = 0.4, 27 boys and 25 girls) participated in Stage 4. They completed the ASTEROID stereotest (version 0.938) and other vision tests in the context of a larger validation study.

#### Procedures

Ethical procedures, data collection procedures and outcome measures were the same as in Stage 1.

#### Impact of PPI

4% of the children showed a poor understanding of the ASTEROID task (defined as less than 80% correct on the practice trials) after we implemented the shuffle animation and the frame cue in the practice trials, compared to 39% before the changes were made. The proportion of correct practice trials significantly increased from an average of 0.76 in the pre-measure to 0.97 in the post-measure (Welch two-sample t-test: t(231)=14.29, p<.001). However, we observed a similar average number of verbal instructions that needed to be given (mean in both pre and post-measures = 10; Welch two-sample t-test: t(77)=0.81, p =.42).

## Study 2: Instructions

### Stage 1: Pre-measure

The post-measure data of Study 1 served as the pre-measure data of Study 2.

### Stage 2: PPI sessions

#### People involved

122 children and adults were involved in the PPI sessions of Study 2. Children were between 1 and 12 years old (median age = 4, IQR = 4). 37 of them were girls and 24 of them were boys. Age and gender was not recorded for the adults. The children attended local science and history museums in Newcastle-upon-Tyne (United Kingdom) in February 2017 and were invited to join a PPI session for 5-10 minutes. Adults were either recruited via our research volunteer pool and registered to attend a session at Newcastle University or they attended a drop-in session at a public venue (Newcastle University Medical School foyer or local museum). Children were thanked for their feedback with a ‘Junior Scientist’ certificate and a sticker.

#### Level and nature of involvement

The public was involved through eight PPI sessions (Table 2). All people involved were given the opportunity to try out ASTEROID (version 0.938-0.94) in drop-in sessions similar to those described in Study 1. Children under 12 were asked for individual feedback and engagement ratings. Sessions with adults were set up to obtain verbal reports on understanding the ASTEROID stereotest.

**Table 2.**
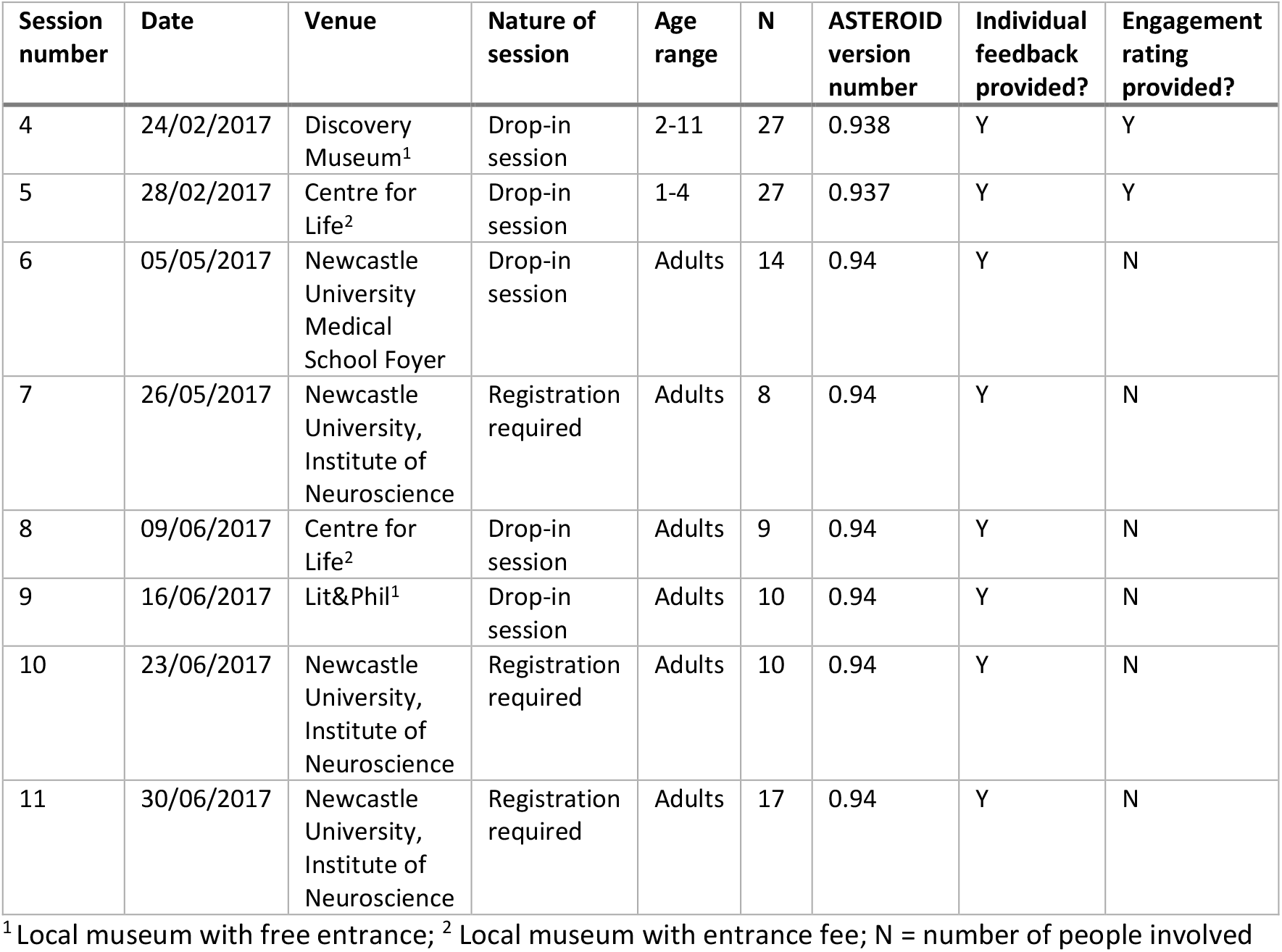
Overview of PPI sessions for Study 2

#### Outcome measures

The outcome measures were the same as in Study 1.

#### Outcomes of PPI

Adults described the target as ‘a square’, ‘centre that sort of sticks out’, ‘popping out and behind’. The outline of the square seems to become more difficult to see at lower disparities when adults described it as ‘a circle appears’, ‘something looks off’, ‘I can see 3D without square’ and even ‘going with the gut’. We used these variety of ways to describe the 3D target to the children but observed that children find it difficult to understand any verbal description of a 3D target. However, once they have seen the target in 3D for the first time, they understand what they need to look out for in the subsequent trials.

### Stage 3: Implementing changes

#### Methods

Just as in Study 1, feedback from Stage 2 was discussed at a cross-disciplinary meeting and consensus changes were implemented in a new version of ASTEROID.

#### Results

We decided to provide the children with a visual and tactile aid to explain them what our 3D target looks like. We therefore made a cardboard demo of an ASTEROID trial. The demo show a print screen of a trial as the background. In the bottom right location, a square with the same dot pattern as the background was glued on top of the background with a 2mm cardboard layer in between (Figure 5).

**Figure 5.**
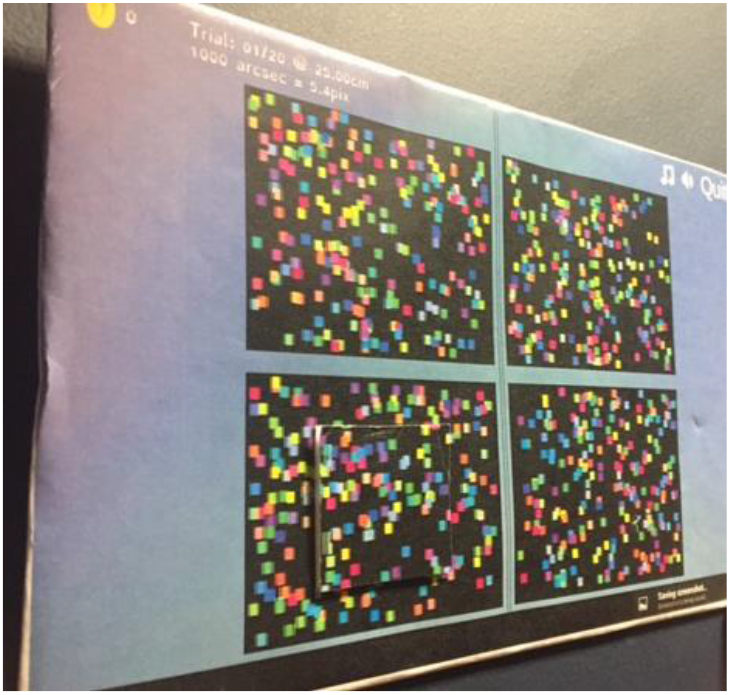
Cardboard demo added after the second Public and Patient Involvement round.

### Stage 4: Post-measure

#### Participants

53 participants between 2 and 40 years old (median age = 10.8, IQR = 5.1, 17 boys and 36 girls) participated in Stage 4. They completed the ASTEROID stereotest (version 0.940) with additional cardboard instructions in the context of a larger validation study.

#### Procedures

Ethical procedures, data collection procedures and outcome measures were the same as in Stage 1.

#### Impact of PPI

6% of the participants showed a poor understanding of the ASTEROID task (defined as less than 80% correct on the practice trials) after we implemented the cardboard instructions compared to 4% without the cardboard instructions. There was no significant increase in average proportion of correct practice trials between pre- and post-measure (mean proportion correct in pre- and post-measure = 0.97; Welch two-sample t-test: t(92)=-0.02, p=.98). However, we found a significant decrease in the average number of additional verbal instructions given from 10 to 8 (Welch two-sample t-test: t(103)=3.72, p<.001). An age difference between the pre- and post-measure might possibly confound this effect, we therefore rerun our analyses including only subjects younger than 9 years old (n=22, median age = 6.3, IQR = 3.4, 8 boys and 14 girls). This indeed resulted in an insignificant difference in the number of additional verbal instructions given (Welch Two sample t-test: t(31)=0.83, p=.41).

## Study 3: Engagement with ASTEROID

Our primary aim was to increase children’s understanding of ASTEROID, however ASTEROID score can only reflect stereo ability if thresholds are not inflated by poor motivation. ASTEROID is therefore embedded in a game and different game themes are available to engage children with different interests. We monitored engagement levels during the development of ASTEROID.

### Methods

Five groups of children and adults were involved in evaluating engagement (Table 3). The groups are described in detail above. Group 4 was a subset of the children and adults involved in Stage 2 of Study 2 (session 4-5). Different outcome measures were collected from different groups (see Table 3). We monitored the number of encouragements given by the researchers during game play, such as ‘Keep going’, ‘Well done’, ‘Just have a go’, etc. In other groups, children rated their level of enjoyment on a smiley face rating scale with five levels (Figure 6). Their ratings were converted to scores between 1 (saddest face) and 5 (happiest face).

**Table 3.**
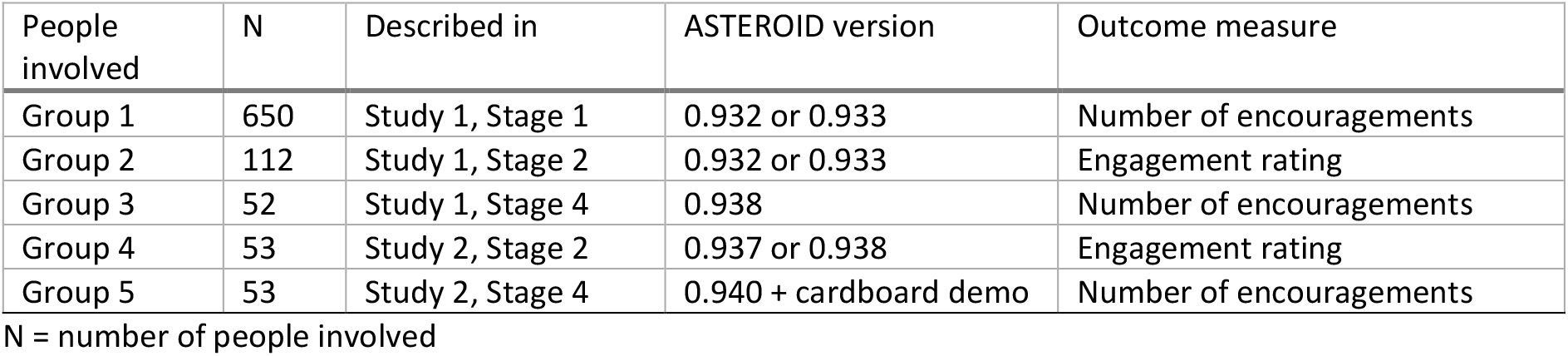
Groups of people involved in evaluating engagement with ASTEROID

| People involved | N | Described in | ASTEROID version | Outcome measure |
| --- | --- | --- | --- | --- |
| Group 1 | 650 | Study 1, Stage 1 | 0.932 or 0.933 | Number of encouragements |
| Group 2 | 112 | Study 1, Stage 2 | 0.932 or 0.933 | Engagement rating |
| Group 3 | 52 | Study 1, Stage 4 | 0.938 | Number of encouragements |
| Group 4 | 53 | Study 2, Stage 2 | 0.937 or 0.938 | Engagement rating |
| Group 5 | 53 | Study 2, Stage 4 | 0.940 + cardboard demo | Number of encouragements |

N = number of people involved

**Figure 6.**
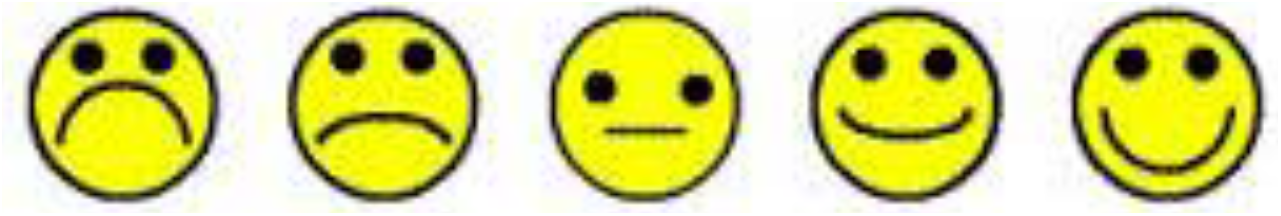
Rating scale for engagement with ASTEROID. The scale was accompanied by the question: Did you enjoy playing the Asteroid game?

### Results

ASTEROID was rated as a fun game by the children with an average rating of 4.8 in Study 1 and 4.7 in Study 2. The implementation of the frame and the shuffle animation did not change the rating (Welch two-sample t-test: t(68) = 0.91, p = 0.36). Although engagement was not the primary aim of the PPI sessions, occasionally positive engagement with ASTEROID was spontaneously noted by the research assistants, for instance ‘Really enjoyed it, laughed at triangle/prize coming up’, ‘Happy to play’, ‘Liked simple [game] but preferred chicken story better’. We found a significant increase of average number of encouragements from 9.6 to 13.8 after we implemented the shuffle animation and the frame cue (Welch two-sample t-test: t(64)=8.25, p<.001). This indicated that children needed more encouragement and ASTEROID was less engaging after Study 1, possibly because of the additional time occupied by the shuffle animation. However, the average number of encouragements significantly reduced to 8.0 after we added the cardboard demo (Welch two sample t-test: t(103)=8.32, p<.001).

## Discussion

Our aim was to improve children’s understanding of the instructions of a new stereotest that measures 3D vision, ASTEROID. Through two PPI studies, we consulted 233 children and adults, an unprecedented number in studies with children. The insights based on their feedback were extremely valuable for the development team and we made three major changes in subsequent versions of ASTEROID: (1) we replaced the colour cue by a frame cue; (2) we added a shuffle animation to explain that the target could appear in any location on the next trial; (3) we made a visual and tactile cardboard demo to explain to the children what target they were looking for in ASTEROID.

The involvement of the public and the changes we made to ASTEROID following their feedback had major impact on the level of understanding of ASTEROID. We noticed a substantial increase in the average proportion of practice trials answered correctly after implementing the frame cue and the shuffle animation. In addition, the percentage of children that showed a poor understanding of ASTEROID (< 80% correct on practice trials) reduced. The impact on the number of additional verbal instructions given to the child during ASTEROID is less straightforward. We observed a slight increase after making the first set of changes, but this decreased again after we included the cardboard demo. This seems to indicate that the cardboard demo was able to replace some of the verbal instructions, while the frame cue and shuffle animation were not. Given the nature of the problem that these changes were trying to solve and the type of verbal instructions given, this is not that surprising. Most verbal instructions were variations on what target to look out for: ‘Tap the one that is different’, ‘Which one is sticking out’, ‘Do any look like they are popping out’? The cardboard demo was included because children had difficulties understanding any verbal descriptions of the 3D target. So, including a visual and tactile aid removed the need for additional verbal instructions. On the other hand, the frame cue and the shuffle animation were solving rather different problems, not directly related to the verbal instructions given.

Our study provides a major contribution to an evidence pool of involvement of children and young people in research. With typical sample sizes of 5-20 children or young people ^17–19^, our sample of 233 people (of which 165 were under 12) is a major step forward in hearing a wide range of opinions. In addition, our study is the first to involve children as young as 1 year old. 30% of our sample are children of 3 years old or younger. Instead of asking parents’ opinions as is commonly done with children that age ^2,5^, we describe methods to successfully gain opinions directly from very young children: using smiley face rating scales for engagement level and using observations and simple questions to investigate understanding of the task. For more in-depth knowledge on the reasoning of solving an ASTEROID trial, we relied on verbal reports from adults unfamiliar with 3D computer tablets. A third strength of our study is the quantitative way in which we measured the impact of our PPI through pre- and post-measures, demonstrating a positive and measurable effect of PPI on research.

The impact of our PPI was possibly limited by the type of people we involved. By running most of our PPI session in public places and through drop-in sessions, we aimed to lower the barriers for involvement. However, most children involved were visiting a local science or history museum with their parents. Museum visitors do not necessarily reflect general demographics. In addition, one of the museum (Centre for Life) has an entrance fee of £11 for adults and £6.50 for children between 5 and 17. This probably caused an underrepresentation of children from lower socio-economic backgrounds. A second limitation of our study is that the median age of participants in our post-measure of Study 2 (median age = 10.8) was higher than in our pre-measure (median age = 5). Our analyses on a subsample of the younger children indeed suggests that increased understanding was mediated by increased age in our sample rather than the cardboard change we have implemented. A larger sample of younger children is needed to confirm this.

We learned the following lessons from our PPI sessions that might be useful for other researchers.

- The drop-in nature of our sessions required very little commitment of the children and the parents, which lowered the barrier for involvement.
- Besides ASTEROID we brought a range of activities and games related to vision and the brain to engage the public, parents and siblings. This certainly helped to attract and engage people.
- We set up our stall/table with activities in a central place in the venue with high volume of passage rather than opting for a quiet separate room as is usually preferred for research in public places.
- The experience of our team in working with children – both clinically and in schools – was extremely valuable. They were able to make children feel at ease, bring over their enthusiasm and be sensitive to a child’s individual needs.

In conclusion, children’s contributions have measurably impacted on the development of ASTEROID, a new stereotest for children. By increasing accessibility and through creative methods we gathered feedback from 165 children between 1 and 12 years old and 68 older children and adults. The changes implemented following their feedback significantly improved our stereotest. Our approach can be inspirational for future researchers and contribute to an evidence pool of good-practice in involvement of children and young people in research.

## Data Availability

Data will be available after publication in journal here: 10.6084/m9.figshare.8345570 and 10.6084/m9.figshare.8345573
Analyses code will be available here: 10.6084/m9.figshare.8378582

## Acknowledgements

We would like to thank Adam O’Neill for administrative support, the orthoptists from Newcastle Eye Clinic for their feedback and assistance in data collection, all children and parents for taking part, our patient panel for their advice at various stages of the research, and all staff of schools, nurseries, and museums in Newcastle-upon-Tyne for hosting and supporting us.

## Conflicts of interests

Jenny CA Read is a consultant for Magic Leap. The ASTEROID vision test has recently been licensed to a company. The other authors declare no other conflicts of interest.

## Funding details

This manuscript presents independent research commissioned by the Health Innovation Challenge Fund (HICF-R8-442, WT102565/z/13/z), a parallel funding partnership between the Wellcome Trust and the Department of Health. The views expressed in this manuscript are those of the authors and not necessarily those of the Wellcome Trust or the Department of Health.

## Notes

### Author Declarations

All relevant ethical guidelines have been followed and any necessary IRB and/or ethics committee approvals have been obtained.

Any clinical trials involved have been registered with an ICMJE-approved registry such as ClinicalTrials.gov and the trial ID is included in the manuscript.

